# Left ventricular global longitudinal strain as a parameter of mild myocardial dysfunction in athletes after COVID-19

**DOI:** 10.1101/2023.03.14.23287258

**Authors:** J Schellenberg, M Ahathaller, L Matits, J Kirsten, J Kersten, JM Steinacker

**Author notes:** Corresponding author: Jana Schellenberg, University Ulm Hospital, Division of Sports- and Rehabilitation Medicine, Leimgrubenweg 14, 89075 Ulm, Germany. Address of the authors: University Ulm Hospital, Division of Sports- and Rehabilitation Medicine, Leimgrubenweg 14, 89075 Ulm, Germany.

## Abstract

**Background:** Whether impaired left ventricular (LV) function contributes to persistent cardiopulmonary symptoms or decreased exercise capacity after COVID-19 remains unclear. The aim of this prospective study was to determine differences in LV global longitudinal strain (GLS) between athletes who did not have a history of LV dysfunction but had a positive COVID-19 test (PCAt) and healthy control (CON) athletes and relate them to symptoms during COVID-19.

**Methods:** We performed 151 transthoracic echocardiographies in our high-performance laboratory. GLS was determined in four-, two-, and three-chamber views and assessed offline by a blinded investigator in 88 PCAt (35% women) at a median of two months after COVID-19 who trained at least three times per week with more than 20 MET per week and 52 CONs from the German national squad (38% women).

**Results:** GLS was significantly lower (GLS -18.53±1.94% vs. -19.94±1.42%, p<0.001) and diastolic function significantly reduced (E/A 1.54±0.52 vs. 1.66±0.43, p=0.020; E’l 0.15±0.04 vs. 0.17±0.04, p=0.009; E/E’l 5.74±1.74 vs. 5.22±1.36, p=0.024) in PCAt. There was no association between GLS and acute symptoms like resting dyspnea, exertional dyspnea during or after COVID-19, palpitations, chest pain or increased resting heart rate. However, there was a trend toward lower GLS in PCAt with subjectively perceived performance limitation (p=0.054).

**Conclusions:** In a cohort of athletes at a median two months after COVID-19, significantly lower GLS and diastolic function were observed, suggesting mild myocardial dysfunction. GLS could be used as a screening element during return-to-sport examinations.

## Introduction

Coronavirus Disease 2019 (COVID-19) is a systemic viral infection caused by Severe Acute Respiratory Syndrome-Coronavirus-2 (SARS-CoV-2) that primarily affects the respiratory system but can also cause myocardial damage (1-3). Even supposedly healthy individuals and athletes may be limited by COVID-19 despite a normally good state of health and fitness. Studies of elite athletes have shown that infection is often mild (46-82%) or asymptomatic (16-58%) (4-7). The most commonly reported symptoms are fever, headache, limb and muscle pain, flu symptoms, fatigue and dyspnea (4, 8, 9). In rare cases (1-3%), myocarditis may occur (10, 11). Most athletes can return to competitive and amateur sports after a training break adapted to the existing symptoms and, if necessary, a return-to-sport examination (12-14). However, having passed through COVID-19 does not necessarily imply complete recovery to the original health or performance status. Symptoms may persist for weeks and months after symptomatic as well as asymptomatic disease progression or reappear with latency (15). Approximately 20-30% of SARS-CoV-2 positive patients in the normal population (16, 17) and probably fewer athletes (1.2-40%) still have symptoms after acute infection (18, 19). Fatigue, neurocognitive impairment, exertional dyspnea (20-30%), exertional and non-exertional chest pain or palpitations (16%) may arise or persist after COVID-19 (13, 17, 20). Acute as well as persistent symptoms can lead to athlete performance loss and even career termination if athletes are unable to return to their pre-COVID-19 performance.

In this context, impaired left ventricular (LV) function could contribute to persistent symptoms and reduced performance. However, larger multicenter studies of return to sport in competitive athletes suggest cardiac involvement in only a few cases (4, 10, 11, 21). If cardiac symptoms exist during and/or after infection, a return-to-sport examination with echocardiography should be performed (13). Here, LV function can be assessed with speckle-tracking echocardiography (STE) in addition to conventional echocardiographic indices (12). Individual studies demonstrated reduced LV global longitudinal strain (GLS) with preserved Ejection Fraction (pEF) in the setting of acute SARS-CoV-2 infection in hospitalized patients regardless of infection severity (22-24) and in patients recovered from COVID-19 (25-27). There are limited data on changes in LV GLS in athletes after COVID-19. In two studies, GLS was not altered after COVID-19 (28, 29).

The aim of this prospective study was, first, to determine differences in LV GLS between athletes who had no history of LV dysfunction but had a positive COVID-19 test (PCAt) and healthy control athletes (CON). Second, we investigated whether there was an association between GLS and symptoms during COVID-19 to identify athletes in need of more targeted follow-up.

## Methods

### Study population

151 competitive athletes presenting to the Ulm Clinic for Sports and Rehabilitation Medicine underwent transthoracic echocardiography in this prospective single-center cohort study between June 2020 and November 2021. For our evaluation we were able to include 140 examinations: 88 athletes with history of a positive COVID-19 test (PCAt) at a median of 2.00 months (25% quantile=1.00 months, 75% quantile=5.00 months) after COVID-19 and 52 healthy athletes from the German national squad as control group (CON) presenting for annual pre-participation screenings. Inclusion criteria for PCAt were: ≥ 18 years of age, training at least 3 times per week with more than 20 metabolic equivalents of task (MET) per week, positive SARS-CoV-2 PCR test or antibody detection with additional typical symptoms. Exclusion criteria for both groups were: acute or chronic medical conditions that precluded the planned physical examination, acute SARS-CoV-2 infection, refusal of peripheral venous blood sampling, inadequate German language skills and withdrawal from study participation. Fifty-seven PCAt (65%) responded to questionnaires about symptoms and complaints during COVID-19, which we included in our analysis. Participants provided written informed consent after being instructed of the study procedures. The study was conducted in accordance with the Declaration of Helsinki and approved by the local ethics committee of the University of Ulm (EK 408/20).

### Echocardiography

151 echocardiographic examinations were performed using an EPIQ 7 ultrasound system with a phased-array probe X5-1 (Philips GmbH, Hamburg, Germany). GLS and global radial strain (GRS) were determined in apical four-, two- and three-chamber views in the apical, midline, and basal segments. They were determined offline using TomTec postprocessing software (2D Cardiac Performance Analysis, TomTec Imaging Systems, Unterschleissheim, Germany) by an investigator who was blinded to group assignment. The endocardial contour was manually adjusted. Eleven echocardiographs (7.3%) with impaired image quality and in which GLS and GRS could not be determined in all standardized apical views were excluded. Segments were classified as normal based on GLS if regional GLS was ≤-16.0% and abnormal if ≥-16.0%. A selection of 15 images were reviewed a second time by the same investigator and another time by a second blinded investigator to determine intrarater and interrater reliability. The following parameters were collected: end-diastolic volume (EDV), end-systolic volume (ESV), left ventricular mass, left ventricular Ejection Fraction (LV-EF by biplane LV planimetry by Simpson), fractional shortening (FS), GLS, GRS, stroke volume (SV), and resting heart rate (HR). Diastolic function was characterized by E/A ratio, E/E’lateral ratio, E/E’medial ratio, V_max_E, V_max_A and deceleration time (Dec Time).

### Statistical analysis

Statistical analyses were performed using R version 4.1.1 (30). Descriptive data are presented as mean (M) ± standard deviation (SD) or as median and 25% quantile and 75% quantile. Group differences were examined using unpaired-Wilcoxon-Tests. Correlations between GLS and age and BMI were analyzed using the Pearson-correlation coefficient (r) and Spearman’s ρ. Additional analyses comparing the frequency of clinically abnormal GLS values in PCAt and CON were calculated using the phi coefficient. To control for possible confounding variables (BMI, age, sex, HR), linear regression models were performed for the confounding variables separately. A p-value of < 0.050 was considered significant.

## Results

### Cohort characteristics

A total of 88 PCAts and 52 CONs were included in the statistical analysis. The groups did not differ in terms of sex, weight, height, systolic blood pressure or HR. A significant age difference was found between PCAt and CON. BMI and diastolic blood pressure were significantly different between the two groups (Table 1).

**Table 1:** Cohort characteristics subdivided in athletes after COVID-19 (PCAt) and healthy athletes of the German national squad (CON)

|  | PCAt | CON | test statistic | p-value |
| --- | --- | --- | --- | --- |
| **Number** | 88 | 52 |  |  |
| **Sex (female/male)** | 31/57 (35%/65%) | 20/32 (38%/62%) | $\phi=0.04$ | 0.741 |
| **Age (years), median (IQR)** | 29 (22 – 40) | 22 (19 – 28) | W=2655.5 | **<0.010** |
| **Weight (kg), median (IQR)** | 73.3 (64.2 – 84.5) | 72.4 (61.4 – 80.9) | W=2735 | 0.212 |
| **Height (cm), median (IQR)** | 176 (168 – 189) | 179 (171 – 184) | W=3332.5 | 0.4072 |
| **BMI (kg/m<sup>2</sup>), median (IQR)** | 22.8 (21.6 – 25.9) | 22.7 (20.8 – 24.3) | W=2522.5 | **<0.050** |
| **Systolic blood pressure, mmHg, median (IQR)** | 120 (110 – 125) | 120 (110 – 130) | W=2746 | 0.1418 |
| **Diastolic blood pressure, mmHg, median (IQR)** | 80 (70 – 80) | 80 (71.25 – 90) | W=2559 | **0.033** |
| **HR (bpm), median (IQR)** | 62 (55 – 68) | 62 (56 – 68) | W=5709 | 0.6317 |
| **Training volume** | at least three times per week<br>> 20 MET | > three times per week<br>>> 20 MET |  |  |

Abbreviations: W (two-tailed unpaired) Wilcoxon Signed Rank Test or $\phi$ Phi-coefficient. IQR interquartile range BMI body mass index HR heart rate bpm beats per minute MET metabolic equivalents of task

### Echocardiographic parameters

PCAt showed significantly lower GLS values than CON (Figure 1A). In both groups, GLS did not differ between men and women. Positive associations were found between GLS and BMI (ρ=0.171, p=0.025) in the total cohort. Intrarater and interrater reliability with respect to the GLS measure showed high agreement (intrarater: 0.892 [95%CI, 0.593-0.973]; interrater: 0.794 [95%CI, 0.159-0.949]. There was no significant association between GLS and age (ρ=0.127, p=0.093), GLS and the time period between examination and infection (ρ=0.155, p=0.115), GLS and HR (ρ=0.146, p=0.054) or between GLS and systolic (ρ=-0.110, p=0.1646) and diastolic blood pressure (ρ=-0.06, p=0.483).

**Figure 1:**
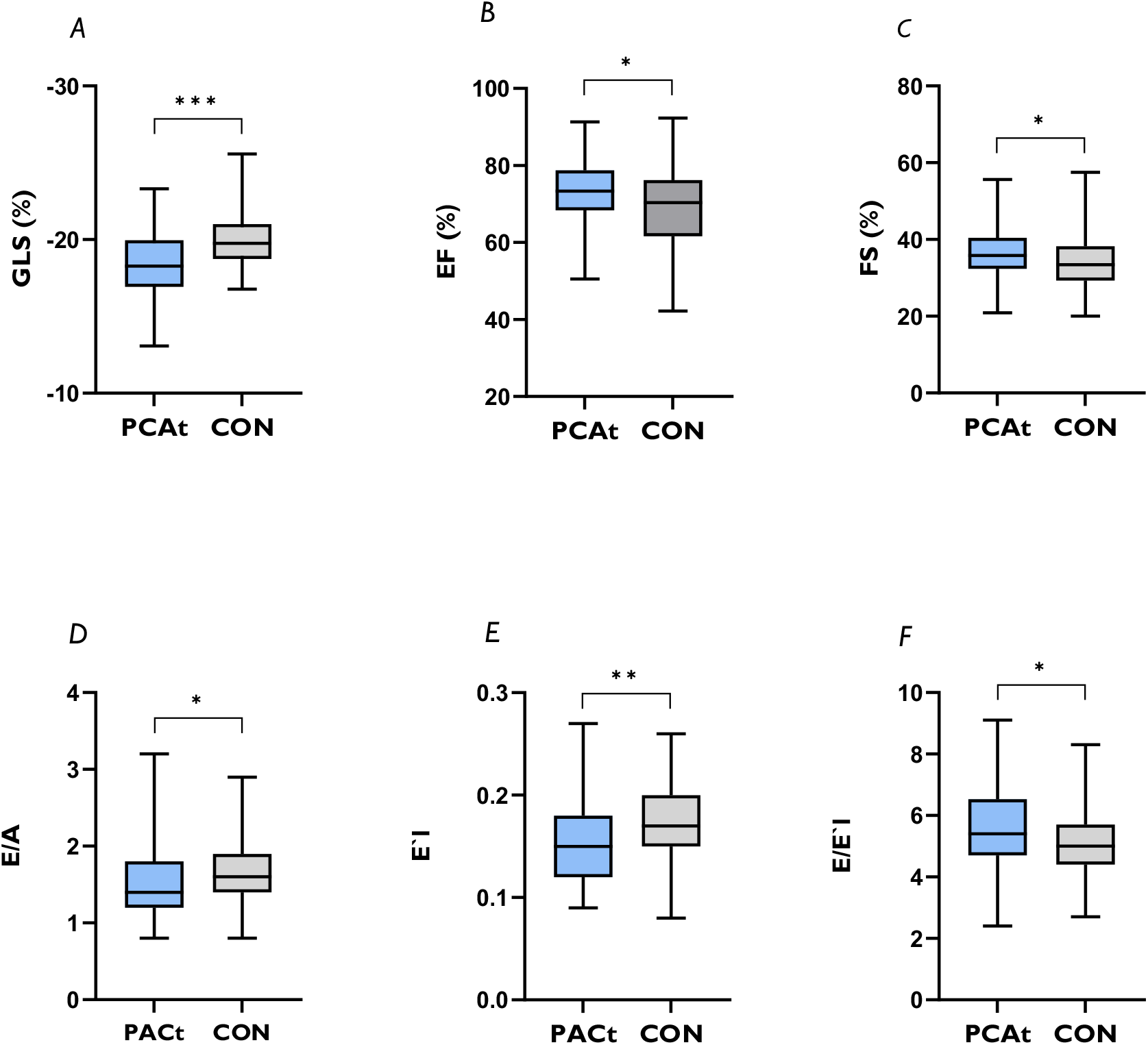
Differences in left ventricular parameters and diastolic function between athletes after COVID-19 (PCAt) and healthy federal squad athletes (CON). A: Global longitudinal strain (GLS) B: Ejection Fraction (EF) C: Fractional shortening (FS) D: E/A ratio E: E’l F: E/E’l. Significant results were presented as follows: * < 0.05 ** < 0.01 *** < 0.001

LV-EF and FS were normal in both groups but significantly higher in PCAt (Figure 1B and 1C). In PCAt, end-systolic volume was significantly smaller and LV mass was significantly reduced (Table S1). Measured values of diastolic function were within the normal range but there was a significant difference for E/A, E’lateral and E/E’lateral (figure 1D, 1E and 1F). The extent of GLS reduction was closely related to E/A-Ratio (p<0.001) and to V_max_E (p<0.001). There were no significant differences in EDV, stroke volume and GRS between PCAt and CON (Table S1). The results of the comparison between PCAt and CON remained when age, HR, sex or BMI were included as control variables.

### Initial symptoms and correlation analysis with GLS and GRS

Symptoms reported during COVID-19 infection were classic symptoms of viral infection: cough (55%), rhinitis (66%), exertional dyspnea (57%), and subjectively perceived reduction in performance (66%) compared with maximal performance before COVID-19. Even after infection, exertional dyspnea persisted in 62% (Table S2). GLS and GRS values did not differ between PCAt who reported symptoms during COVID-19 compared with asymptomatic PCAt. (Table S3 and S4). However, there was a trend toward lower GLS in PCAt with subjectively perceived performance limitations, but it was not significant (W=417.0, p=0.057).

## Discussion

Cardiopulmonary symptoms after COVID-19 have been described, but not the influence of disease symptoms on cardiac function. In this prospective, single-center cohort study, we observed significantly lower GLS values and diastolic function in PCAt, suggesting mild myocardial dysfunction which might lead to small but decisive performance losses in competitive sports.

Acute SARS-CoV-2 infection in hospitalized patients showed reduced GLS with pEF, regardless of the severity of infection (22-24), but the long-term consequences, e.g., in terms of mortality, incidence of heart failure, or eventual recovery, are still unknown. Lower GLS has also been observed in patients with heart failure and pEF (31). There is limited research on GLS in athletes after COVID-19. Fikenzer et al., found no echocardiographic GLS differences between eight infected and four uninfected elite handball players, but magnetic resonance imaging showed mild signs of acute inflammation/edema in all infected athletes (29). However, the study was very small and limited to male athletes. In our study, ten PCAt with reduced GLS and persistent symptoms received MRI. In three of eight MRI examinations myocarditis was revealed, which is consistent with data from the current literature describing a very low rate (1-3%) of myocarditis after COVID-19 (10, 11). Echocardiographic GLS measurement is a commonly available test and may be useful as an additional parameter in selecting athletes with appropriate clinic for limited capacity MRI examinations (12). Another study showed no COVID-19-mediated GLS differences in 107 elite athletes (23% women) with mild symptoms and 107 randomized healthy athletes. (28). However, in a subset of post-COVID athletes with early diastolic septal flattening, there was a relative decrease in GLS in the free wall segments, suggesting a characteristic feature of pericardial constriction. Concerning the LV-EF, however, our results are in line with Lakatos et al. showing increased LV-EF in PCAt compared with CON. Increased LV-EF can result from reduced training volume, since LV-EF is often low normal (32) or even slightly reduced in athletes (28, 33). Here, longitudinal investigations of GLS and LV-EF in detraining would be necessary.

Decreased GLS as an early marker of myocardial dysfunction could potentially contribute to persistent cardiopulmonary symptoms and subjectively perceived performance limitation after COVID-19. We investigated whether symptomatic courses lead to cardiac sequelae but found no differences in GLS or GRS between symptomatic and asymptomatic athletes. Classic symptoms such as cough, rhinitis, sore throat, exertional dyspnea, and subjectively perceived performance limitations were present in more than half of the PCAt, as reported in several studies (4, 8, 9). Even after infection, exertional dyspnea remained in 62% of PCAt, which is significantly more frequent than 20-30% reported in the literature (13, 20). Reasons for this could be: First, athletes might notice small changes in form of exertional dyspnea earlier and more than non-athletes or secondly, it could be that mainly athletes with complaints presented themselves to our department. However, there was a trend toward lower GLS in PCAt with subjectively perceived performance limitation. A direct effect of reduced GLS on performance does not yet exist and should be evaluated in further studies. To our knowledge, this is the first study to evaluate individual symptoms rather than severity of course. Post-COVID patients with significant functional impairment were more likely to have impaired GLS or cardiovascular comorbidities whereas those with mild to moderate functional impairment had no cardiovascular changes (25). Similar results were obtained by Mahajan et al., who described increasing impairment of GLS in mild (13.1%), moderate (44%), and severe (90%) disease (26). These patients were older than our cohort of athletes and had concomitant cardiovascular diseases such as hypertension and diabetes mellitus. Furthermore, they were examined earlier (30-45 days after infection) than ours, so improvement of LV dysfunction cannot be excluded. However, our results argue against a temporal component: There was no significant association between GLS and the time period between examination and infection, which could indicate a possible stable long-term decrease in GLS in PCAt.

In our study, there is no positive correlation between GLS and age in the overall cohort. Zghal et al. found a decrease in GLS with age with no change in LV-EF (34). In the Danish City of Copenhagen Heart Study GLS changed differently with age in men and women, with men having lower mean values and lower reference limits for all exercise parameters. 62% of participants were female and older with an age of 46±16 years compared with our study, which had 45% women and ages of 31.44±12.62 (PCAt) and 24.69±7.89 (CON) years, respectively (35). In our study, GLS did not differ between men and women in both the PCAt and CON groups. Although diastolic blood pressure was significantly different between PCAt and CON, there was no association between GLS and systolic or diastolic blood pressure. Blood pressure levels correlated with GLS (36, 37), and GLS was significantly reduced in patients with hypertension compared with normotensive control subjects (38). Due to the specific athlete clientele, the variance of the blood pressure values is low, which is accompanied by a limited interpretability of the correlation. There was a positive association between GLS and BMI in our study. Impairment of LV-EF and GLS by overweight and obesity is also known from other studies (39-41). Parameters of diastolic function were within the normal range in our studied athletes but were significantly reduced in PCAt. Some authors indicate that diastolic function is normal or decreased in athletes (42, 43) but may also be supranormal in trained endurance athletes compared with untrained individuals (44). According to the study by Galderisi et al., the extent of GLS is closely related to diastolic function (43). We also demonstrated a correlation of GLS with E/A ratio and V_max_E. Thus, routine determination of diastolic function may be an initial clue to possible LV dysfunction and should be followed by determination of GLS at the latest in case of abnormalities.

### Strengths and Limitations

This study is limited by the cross-sectional design, as no prior strain values of PCAt and CON were available. Therefore, it cannot be excluded that reduced strain values did exist preliminary or are due so sport-related adjustments. Because of the nature of this study, the time after SARS-CoV-2 infection differed between subjects, and heart function may have already improved or worsened during infection perhaps as a result of detraining. However, there was no significant correlation between GLS and the time between examination and infection. The clearly defined population of study participants, consisting of athletes, limits the generalizability of the results for the general population, but on the other hand allows an assessment of a specific group. The difference in training volume between the two groups may affect our results because GLS is sport-dependent. However, even when the groups differed significantly in age and diastolic blood pressure, no significant association between GLS and age and GLS and diastolic blood pressure was found. Although the GLS determination can be software and investigator experience dependent, our results show high intrarater and interrater reliability. It should be emphasized that we achieved a meaningful case number of athletes for a single-center study. Finally, although an association between COVID-19 and the occurrence of pathological examination findings up to myocarditis is suggested, direct evidence is still lacking.

## Conclusion

Significantly lower GLS and diastolic function in PCAt compared to healthy peers suggests mild myocardial dysfunction after COVID-19, potentially contributing to decreased training and competition performance. When it comes to success and failure, winning medals or top placings, small differences in performance can be decisive. Therefore, myocardial damage, in addition to factors such as lack of training can be detrimental to success. Therefore, determination of GLS as a screening element for early detection of left ventricular dysfunction could be part of the return-to-sport examination. Long-term observations in athletes as well as in the general population are needed to evaluate the impact of COVID-19 on cardiac function and performance limitation.

## Data Availability

The data underlying this article will be shared on reasonable request to the corresponding author.

## Supplementary Data

Table S1: Echocardiographic parameters subdivided in athletes after COVID-19 (PCAt) and healthy athletes of the German national squad (CON)

Table S2: Symptoms during COVID-19 in athletes after COVID-19 (PCAt) presented as absolute values and relative frequencies

Table S3: Symptoms during COVID-19 in athletes after COVID-19 (PCAt) in correlation with GLS.

Table S4: Symptoms during COVID-19 in athletes after COVID-19 (PCAt) in correlation with GRS.

## Funding

This work was supported primarily by the University Hospital Ulm and funds from the German Federal Institute for Sport Science, Cologne by resolution of the German Bundestag [ZMVI4-070106/20-23].

## Conflicts of Interest

Nothing to Disclose.

## Data Availability

The data underlying this article will be shared on reasonable request to the corresponding author.

## Author Contributions

J.S. and J.M.S. responsible for conceptualization; M.A. and J.S. performed measurements, L.M. and M.A. analyzed data; J.S., M.A., L.M. and J.M.S. interpreted results of data and examinations; J.S. and L.M. prepared figures; J.S. drafted manuscript; L.M., J.Ki., J.K. and J.M.S. edited and revised manuscript; J.S., L.M., J.K. and J.M.S. approved final version of manuscript.

## Notes

### Competing Interest Statement

The authors have declared no competing interest.

### Author Declarations

Ethics committee University of Ulm (408/20) gave ethical approval for this work.

