## Supplementary Data for "Left ventricular global longitudinal strain as a parameter of mild myocardial dysfunction in athletes after COVID-19"

**Table S1**: Echocardiographic parameters subdivided in athletes after COVID-19 (PCAt) and healthy athletes of the German national squad (CON)

|  | **PCAt** | | | **CON** | | |  | |
| --- | --- | --- | --- | --- | --- | --- | --- | --- |
|  | 25% | Med | 75% | 25% | Med | 75% | W | p-value |
| **EDV (ml)** | 108.00 | 123.00 | 146.00 | 104.00 | 141.50 | 169.00 | 4207.5 | 0.246 |
| **ESV (ml)** | 24.18 | 32.50 | 44.53 | 28.62 | 38.80 | 53.40 | 4488.0 | **0.047*** |
| **LV Mass (g)** | 124.00 | 149.50 | 184.75 | 134.00 | 172.00 | 202.50 | 4457.5 | **0.037*** |
| **LV-EF (%)** | 67.85 | 73.25 | 78.57 | 61.95 | 70.35 | 76.00 | 3130.5 | **0.042*** |
| **FS (%)** | 32.32 | 35.75 | 40.30 | 29.35 | 33.40 | 38.00 | 2949.0 | **0.015*** |
| **GLS (%)** | -19.72 | -18.27 | -17.14 | -20.72 | -19.76 | -18.99 | 2059.5 | **<0.001*** |
| **GRS (%)** | 5.60 | 10.42 | 15.91 | 6.11 | 11.12 | 19.45 | 4020.5 | 0.349 |
| **Stroke Volume (ml)** | 77.60 | 89.10 | 110.00 | 71.85 | 92.30 | 115.00 | 3788.5 | 0.855 |
| **E/A** | 1.20 | 1.40 | 1.80 | 1.40 | 1.60 | 1.90 | 4416.0 | **0.020*** |
| **E`l** | 0.12 | 0.15 | 0.18 | 0.15 | 0.16 | 0.20 | 4057.0 | **0.009*** |
| **E`m** | 0.09 | 0.11 | 0.13 | 0.10 | 0.12 | 0.13 | 3129.5 | 0.414 |
| **E/E´l** | 4.70 | 5.45 | 6.60 | 4.40 | 5.00 | 5.70 | 2629.0 | **0.024*** |
| **E/E´m** | 6.30 | 7.30 | 9.15 | 6.50 | 7.20 | 8.10 | 3585.5 | 0.618 |
| **V_max_ A** | 0.46 | 0.55 | 0.64 | 0.46 | 0.52 | 0.58 | 3078.0 | 0.076 |
| **V_max_ E** | 0.67 | 0.81 | 0.96 | 0.72 | 0.83 | 0.94 | 3974.0 | 0.332 |
| **Dec Time** | 129.00 | 164.00 | 198.00 | 144.25 | 183.50 | 218.50 | 2415.0 | 0.168 |

Abbreviations: 25% quantile, Med median, 75% quantile, EDV end-diastolic volume ESV end-systolic volume, LV mass left ventricular mass, LV-EF left ventricular ejection fraction by Simpson, FS fractional shortening, GLS global longitudinal strain, GRS global longitudinal strain, E/A ratio, E´l, E´m, E/E´l ratio E/E´m ratio, V_max_ A velocity of A wave, V_max_E velocity of E wave, Dec Time Deceleration Time.

**Table S2:** Symptoms during COVID-19 in athletes after COVID-19 (PCAt) presented as absolute values and relative frequencies

| **Symptoms** | **Present** | **Not-present** |
| --- | --- | --- |
| fever | 26 (46%) | 30 (54%) |
| cough | 31 (55%) | 25 (45%) |
| rhinorrhea | 37 (66%) | 19 (34%) |
| sore throat | 33 (59%) | 23 (41%) |
| resting dyspnea | 14 (25%) | 42 (75%) |
| exertional dyspnea during COVID-19 | 32 (57%) | 24 (43%) |
| exertional dyspnea after COVID-19 | 34 (62%) | 21 (38%) |
| palpitations | 20 (36%) | 36 (64%) |
| chest pain | 20 (36%) | 36 (64%) |
| increased resting heart rate | 25 (45%) | 31 (55%) |
| subjective perceived performance limitation | 37 (66%) | 19 (34%) |
| dizziness | 25 (45%) | 30 (55%) |

**Table S3:** Symptoms during COVID-19 in athletes after COVID-19 (PCAt) in correlation with GLS

|  | GLS | | | | | | | |
| --- | --- | --- | --- | --- | --- | --- | --- | --- |
|  | Present Symptoms | | | Not present Symptoms | | |  | |
|  | 25% | Med | 75% | 25% | Med | 75% | W | p-value |
| fever | -19.71 | -17.73 | -16.90 | -19.21 | -17.93 | -17.01 | 372.5 | 0.890 |
| cough | -19.43 | -17.35 | -16.97 | -19.32 | -18.17 | -16.90 | 304.0 | 0.363 |
| rhinitis | -19.58 | -17.83 | -17.06 | -19.25 | -17.83 | -16.90 | 352.0 | 0.731 |
| sore throat | -19.05 | -17.38 | -16.53 | -20.22 | -18.11 | -17.23 | 232.5 | 0.036 |
| resting dyspnea | -19.59 | -17.93 | -16.99 | -19.30 | -17.78 | -16.91 | 302.0 | 0.671 |
| exertional dyspnea during COVID-19 | -19.57 | -17.73 | -17.09 | -19.31 | -18.05 | -16.76 | 357.0 | 0.937 |
| exertional dyspnea after COVID-19 | -19.59 | -17.83 | -17.16 | -19.30 | -18.04 | -16.45 | 359.0 | 0.510 |
| palpitations | -19.29 | -18.05 | -17.30 | -19.33 | -17.58 | -16.76 | 385.0 | 0.435 |
| chest pain | -19.77 | -17.63 | -16.71 | -19.27 | -17.93 | -16.95 | 340.5 | 1.000 |
| increased resting heart rate | -19.64 | -17.93 | -16.78 | -19.28 | -17.78 | -16.95 | 361.5 | 0.986 |
| subjective perceived performance limitation | -18.05 | -17.05 | -16.46 | -19.60 | -18.03 | -17.19 | 417.0 | 0.057 |
| dizziness | -19.76 | -17.83 | -17.05 | -19.29 | -17.83 | -16.89 | 394.5 | 0.585 |

Abbreviations: 25% quantile, Med median, 75% quantile, W (two-tailed unpaired) Wilcoxon Signed Rank Test.

**Table S4:** Symptoms during COVID-19 in athletes after COVID-19 (PCAt) in correlation with GRS

|  | GRS | | | | | | | | |
| --- | --- | --- | --- | --- | --- | --- | --- | --- | --- |
|  | Present Symptoms | | | Not present Symptoms | | | |  | |
|  | 25% | Med | 75% | 25% | Med | 75% | W | | p-value |
| fever | 4.33 | 8.53 | 14.60 | 0.86 | 6.31 | 14.83 | 329.0 | | 0.553 |
| cough | 2.88 | 6.27 | 15.40 | 5.69 | 8.16 | 14.04 | 382.0 | | 0.665 |
| rhinitis | 3.45 | 6.78 | 15.40 | 1.67 | 6.49 | 12.18 | 304.0 | | 0.615 |
| sore throat | 3.67 | 6.38 | 14.90 | 3.26 | 7.74 | 14.53 | 375.0 | | 0.695 |
| resting dyspnea | 6.27 | 12.04 | 17.24 | 0.86 | 6.32 | 14.02 | 195.0 | | 0.096 |
| exertional dyspnea during COVID-19 | 2.91 | 6.64 | 14.93 | 4.01 | 6.84 | 14.50 | 355.0 | | 0.965 |
| exertional dyspnea after COVID-19 | 4.22 | 7.47 | 14.60 | -0.42 | 6.27 | 12.94 | 287.0 | | 0.514 |
| palpitations | 5.01 | 8.06 | 15.54 | -0.69 | 6.43 | 14.12 | 279.0 | | 0.281 |
| chest pain | 2.80 | 5.84 | 15.60 | 3.19 | 8.75 | 14.13 | 368.0 | | 0.625 |
| increased resting heart rate | 3.95 | 6.29 | 15.54 | 3.10 | 7.05 | 14.13 | 364.0 | | 0.952 |
| subjective perceived performance limitation | 0.11 | 6.36 | 14.01 | 4.20 | 6.78 | 15.47 | 291.0 | | 0.671 |
| dizziness | 3.95 | 6.19 | 14.75 | 1.11 | 10.31 | 14.66 | 394.0 | | 0.594 |

Abbreviations: 25% quantile, Med median, 75% quantile, W (two-tailed unpaired) Wilcoxon Signed Rank Test.
